# ASSESSING SYSTEM PERFORMANCE OF MATERNAL AND CHILD HEALTH CARE DURING COVID-19 PANDEMIC IN COMMUNITY HEALTH CENTER- COMORO DILI TIMOR-LESTE

**DOI:** 10.1101/2023.10.23.23297388

**Authors:** Agusta Amaral Lopes, Delfim da Costa Perreira, Domingos Soares, Valente da Silva, Nelson Martins

## Abstract

**Purpose/ Introduction:** Recent Evidences reveal that COVID-19 pandemic caused MCH services interruption world-wide. In Timor-Leste, MCH services is one of service priorities and delivers through 3 tiers of health structures consists of hospitals, CHCs, and HPs. The Country (Timor-Leste) identified its first case of COVID-19 in March and by April 2020, it was lockdown and stringent actions were enforced. During the pandemic COVID-19, the MOH health structures and facilities continued delivers essential health services. However, the strict lockdown and mandatory stay home order had negatively impacted the health system capacities. This study to thoroughly assess the disruption of System Components of MCH Services by interviewed frontline HCWs from Comoro -CHC, in Dili-Timor-Leste.

**Patients and Methods:** The cross-sectional approach with quantitative descriptive method was employed for this study. We employed a modified who six building blocks (service delivery, human resources, drugs and consumables, ICT, Financing, Stewardship) to assess system disruption of MCH services in Comoro CHC due to COVID-19 Pandemic. Sample of 99 participants consisted of medical doctors, midwives, nurses who work at MCH unit pharmacy technicians, unit laboratory technicians, a structured questionnaire was utilized and data analysis was used SPSS.

**Result:** The findings reveal the disruption of all 6 system blocks from MCH services in Comoro-CHC, Dili, Timor-Leste due to COVID-19 Pandemic. Except leadership, statistical tests reveal statistically significant association between interruption of five blocks from MCH services. The service delivery, human resources, drugs and consumables, ICT, Financing, Stewardship.

**Conclusion:** COVID-19 Pandemic Caused disruption of all six health system blocks of MCH Services in Comoro, CHC. For future pandemic preparation plan, the attention must be given to all six-health system blocks to guarantee continue delivery of MCH care in CHC Comoro, Dili, Timor-Leste and a primary health care facility and other similar settings.

## INTRODUCTION

Global context: MCH services delivery during covid-19 pandemic. Maternal and Child Health (MCH) refers to the services provided to women in their child bearing and children. [1] [2]. MCH had been placed as a high priority in the most of the countries health systems. The introduction of proper MCH strategy and interventions through antenatal care (ANC), institutional delivery, and post-partum care can prevent maternal and mortality and other related complications. [3] Before COVID-19 Pandemic occurs, there was declined of maternal deaths worldwide of nearly half (44%) since 1990, and a significantly increased use of maternity services. [4] However, the recent data reveal that about 287 000 women died during and following pregnancy and childbirth in 2020. Similarly, according the recent estimation by UN Inter-agency Group for Child Mortality Estimation, there were 5 million children died before their fifth birthday in 2021 alone. [5]

The most common causes that contributed directly the injury and death of women during childbearing were extensive hemorrhagic, infection, hypertension, unsafe abortion, and obstructive labor. There were also indirect causes such as Anemia, malaria, and heart disease. [6] These causes can be prevented with quickly assisted by skilled HCWs working in the well-established MCH system. However, for health system to function well, the attention and resources must be given the its six components such as stewardship; services delivery; human resources; essential medicines & consumables; Information, communication, technology; and financing. [7]

The interruption from one of the above components will compromise health system capacities to deliver quality of MCH Care. So far there are little researches commissioning to examine the disruption of health system components (blocks) of MCH services in primary health care facilities during major disease pandemics. The COVID-19 pandemic has had a disastrous effect on the system for delivery health care of people on a global scale, but pregnant women face particular challenges [8] The public health system was at increased risk of reduced efficiency include MCH services. [9]

Pandemic created obstacles for women and child worldwide to accessing MCH services, including restrictions, transport challenges, and anxiety over possibly being exposed to coronavirus. The movement restriction has made it difficult for many pregnant women to reach health care facilities. Even those who managed to reach health facilities have reported not receiving timely care. [8] The disruption in access MCH services such as routine check-ups, sonography tests, scans, institutional deliveries, postnatal check-ups, vaccination of children, etc., has led to increased suffering of pregnant women and children. [3].

Researches from many developing countries report on the disruption of MCH services during COVID-19 Pandemic and its stringent measures. India estimates, antenatal care has dropped by 20 to 30 percent, institutional delivery by 30 to 43.25 percent and postnatal care by 20 to 30 percent. Around 25-30 percent fall in common doses of vaccines given to children. [3] From Pakistan, Faran report on the Reduced utilization for all RMNCH indicators with the highest reduction (∼82%) was among children aged < 5 years, who were treated for pneumonia during the first wave of pandemic. The number of caesarean sections dropped by 57%, followed by institutional deliveries and first postnatal visit (37% each). [10]

Studies from Ethiopia reveal services disruption of MCH services and declining in health facilities visits for Family Planning, sick under 5 child visits (225.0 to 139.8/ month, p=0.007), Antenatal and postnatal care visits, facility delivery rates [11] [12] Similarly, study from Guyana, revealed significant decreases in utilizing all essential health services across the 41 health centres of Region 4. [13] A rapid review carried out on 20 March 2022 on postnatal care (PNC) services availability and utilization during the COVID-19 era in sub-Saharan Africa revealed that there were significant declines in the availability and utilization of PNC services during and after the COVID-19 lockdown. [14]

In response to COVID-19 pandemic there are increasing call highlighted the importance of health system preparedness and better response with special attention given to vulnerable people like pregnant women and newborns to minimize lag in service delivery. [8] [9] It was argue that while priorities were given to fight COVID-19, there are important to quickly reinstate routine services with key areas include continued provision of family planning services along with uninterrupted immunization campaigns and routine maternal and child services. [10] For most developing countries those routine services could only be effectively implemented through the primary health facilities located closed to the community. Experiences from Indonesian suggest that during the COVID-19 pandemic, Community Health centers (CHCs) a primary health care facility is required to play a role in preventing, detection, and handling COVID-19 with optimal, while continued delivering MCH essential Services. [15] [16]

However, Horton R, 2021 argue that the pandemic has created further obstacles in the health system to provide MCH services by healthcare facilities. [17] According to Ani KS and friends, during the pandemic, healthcare workers, equipment, and healthcare facilities have been transformed into COVID hospitals to cope with the rising numbers of infected people, leaving little space for the healthcare of pregnant women [9]. Bisht R, Sarma J., 2020 further augmented that women who managed to reach health facilities have reported not receiving timely care due to shortage of health care workers in some health centers, because many were engaged in treating COVID-19 patients, while in other centers maternal health services were cut despite being classified as essential service. [18].

### Timor Context

Timor Leste (TL) reported good improvement in the indicators of Maternal and Child Health during the two decades of Independence. Despite this, the rates are still considered high compared to other Asian countries with maternal mortality rate of 218 per 100,000 live births and under-five mortality of 41 deaths per 1,000 live births (TDHS, 2016). The MCH services has been become one of the priorities services and delivered through 3 tiers of health systems consists of hospitals, community health centers, health posts and through primary health care strategies such as SISCa, SnF, and Villages Midwives. In 2020, in responds to COVID-19, Ministry of Health issued a policy to guide the continued implementation of essentials health services, including MCH care. Several activities in MCH services must be carried out, were including the first and third trimester pregnancy checks, normal deliveries for non-COVID-19 cases, routine postpartum and family planning services, first postpartum visits, neonatal services, and immunization. (Politica, 2020).

TL reported it first COVID-19 cases in March, 2020 and by October 27 2021, it registered 19.785 cases with 121 deaths. [19] TL health system was still young and not ready to deal with COVID-19 Pandemic. [20] Despite of its huge challenges, the MOH structures and health facilities continue to deliver essential health services during COVID-19 Pandemic. However, the strict lockdown and mandatory stay home measures have negatively impacted essential services delivery. [21] Two nationals’ studies focusing on services delivery component of MCH during COVID-19 pandemic reveal contradictory results of the disruption of MCH services. [22] [23]

So far there is no study commissioning to assess health system readiness for delivering MCH services in Timor-Leste. We conducted this study to thoroughly assessed system performances for delivering MCH services during COVID-19 Pandemic in a Community Health Centers, Comoro Dili, Timor-Leste

## MATERIAL AND METHODS

The cross-sectional approach with quantitative descriptive method was employed to assess the system performance of MCH services. We employ a modified WHO six building blocks in assessing performance of health systems, [7] to assess the system performance of Comoro CHC in Dili in delivering MCH services during pandemic COVID-19.

For this study, we questioned health staff’s opinion on the availability each system components (stewardship, services delivery, human resources, Drugs & Consumables, ICT and Financing) in the provision of MCH Care during COVID-19 pandemic (Table-1)

**Table-1:** Operational Definition Variables and Measurement Tools.

| No | Variables | Operational Definition | Measurement Tools | Scale | Category Response |
| --- | --- | --- | --- | --- | --- |
| **Independent** |  |  |  |  |  |
| 1. | Service Delivery | Delivery services MCH during COVID-19 Pandemic | Questioner | Nominal | 1. Yes. Services available<br>2. No. Services was not available<br>3. |
| 2. | Health Staffs | Availability of Health Staffs to Delivery MCH services during | Questioner | Nominal | 1. Yes. Available<br>2. No. Not Available |
| 3. | Information, Communication, Technology (ICT) | Availability of ICT for MCH services | Questioner | Nominal | 1. Yes. Available<br>2. No. Not Available |
| 4. | Essential Medicine and Medical Equipment | The availability of essential medicine and Equipment for MCH care | Questioner | Nominal | 1. Yes. Available<br>2. No. Not Availbale |
| 5. | Financing | Availability of Funding for MCH services | Questioner | Nominal | 1. Yes. Available<br>2. No. Not Available |
| 6. | Stewarship | Stewarship for MCH services provided | Questioner | Nominal | 1. Yes. Available<br>2. No. Not Available |
| **Dependent** |  |  |  |  |  |
| 7. | MCH Care | Total MCH visits to health facilities during COVID-19 Pandemic | Questioner | Nominal | 1. Yes. There are visits<br>2. No. No visits |

This study was conducted from November 2021 to April 2022.

A sample of 99 participants enrolled for the interview. The interviewees consisted of medical doctors, midwives, nurses who work at maternal and child Health (MCH) unit; pharmacy technicians who work in the pharmacy unit; and laboratory technicians who work in laboratories. N*on-probability sampling Consecutive Sampling* was utilized to identified and enrolled participants.

### Study Setting

This study is conducting in the one of main community health centers in Dili Municipality, Timor-Leste. Timor-Leste is small countries divided into 12 Municipalities, and 1 especial region of Oecusse with Population of 1.321 million. [24] The Comoro CHC is one of the main health centers with in-patients care with 12 beds, an emergency room, maternity clinic, laboratory and out-patients care. It located in Comoro CHC has served a population of 159,773 people with an area of 33.12 km². The working area of the Comoro health center is located in the Posto Administrativo Dom Alexio area, Dili Municipality, Timor-Leste, which has 7 villages.

The Government health facilities in the area consist of one community health center (Comoro CHC) and eight health posts. There are 102 employees working at the Comoro Community Health Center and 45 employees working at the Health Post. Some of these employees have the status of civil servants and there are also contract employees who are responsible for organizing and implementing health programs on a routine basis and in emergency situations. About 14 priorities programs, including MCH care were implemented regularly in this health center, see - table-1.

#### Data Collection

A structured questionnaire was employed and purpose sampling method was utilized to enroll participants. The interviewed results were entering into excel spreadsheet and transferred to SPSS for statistical analysis. The structure questioners composed of 21 questions based on each research variables and category response listed in table-1 below), were administered to collect the response from respondents.

#### Data Analysis

The results were entering into excel spreadsheet and transferred to SPSS (version) for statistical analysis.

1. Univariable analysis was carried out to determine the characteristics of the sample descriptively with a frequency distribution
2. Bivariable Analysis was employed to determine the influence between two variables, namely the independent variable and the dependent variable without control, to assess whether the influence of the independent variable and the dependent variable is statistically significant using the Chi-Square test with the degree of significance used is 95% and the p value with the level significance is p_J0.25.

## RESULTS

### Respondent Characteristics

We have successfully interviewed 94 participants compose of midwives, Nurses, Medical Doctors, Pharmacist and Medical Scientists working in supporting MCH services in CHC Comoro during COVID-19 Pandemic. Information from Table-2 below, reveal that about 70% of respondents were temporary contract and voluntary staffs; with 56,4 % has less than 5 years working experiences, and 90.4 % with them ages between 24-36 years old. Most respondents (77.7%) was female and was tasked to deliver routine MCH services in Comoro CHC. However, during the COVID-19 Pandemic around 24.5% of them did not delivery MCH services.

**Table-2:** The Essential Health Programs Implemented at Comoro – CHC.

| NO | HEALTH PROGRAMS |
| --- | --- |
| 1 | Maternal and child health; Reproductive health, Family Planning, ANC, PMTCT, MCH/obstetrics, PNC, newborn care, IMCI and Immunization. |
| 2 | Infectious diseases such as: TB, Leprosy, Malaria, STI and HIV/AIDS, Syphilis |
| 3 | Nutrition |
| 4 | Non-communicable diseases such as; Mental Health, Oral Health, Elderly Health |
| 5 | Treatment: adults, and children |
| 6 | Environmental Health |
| 7 | Promotion of school health & wellness |
| 8 | Information systems |
| 9 | Pharmacy |
| 10 | Laboratory |
| 11 | SISCa |
| 12 | SnF/Family Health |
| 13 | Finance |
| 14 | Administration and Logistic |

**Table 2:**
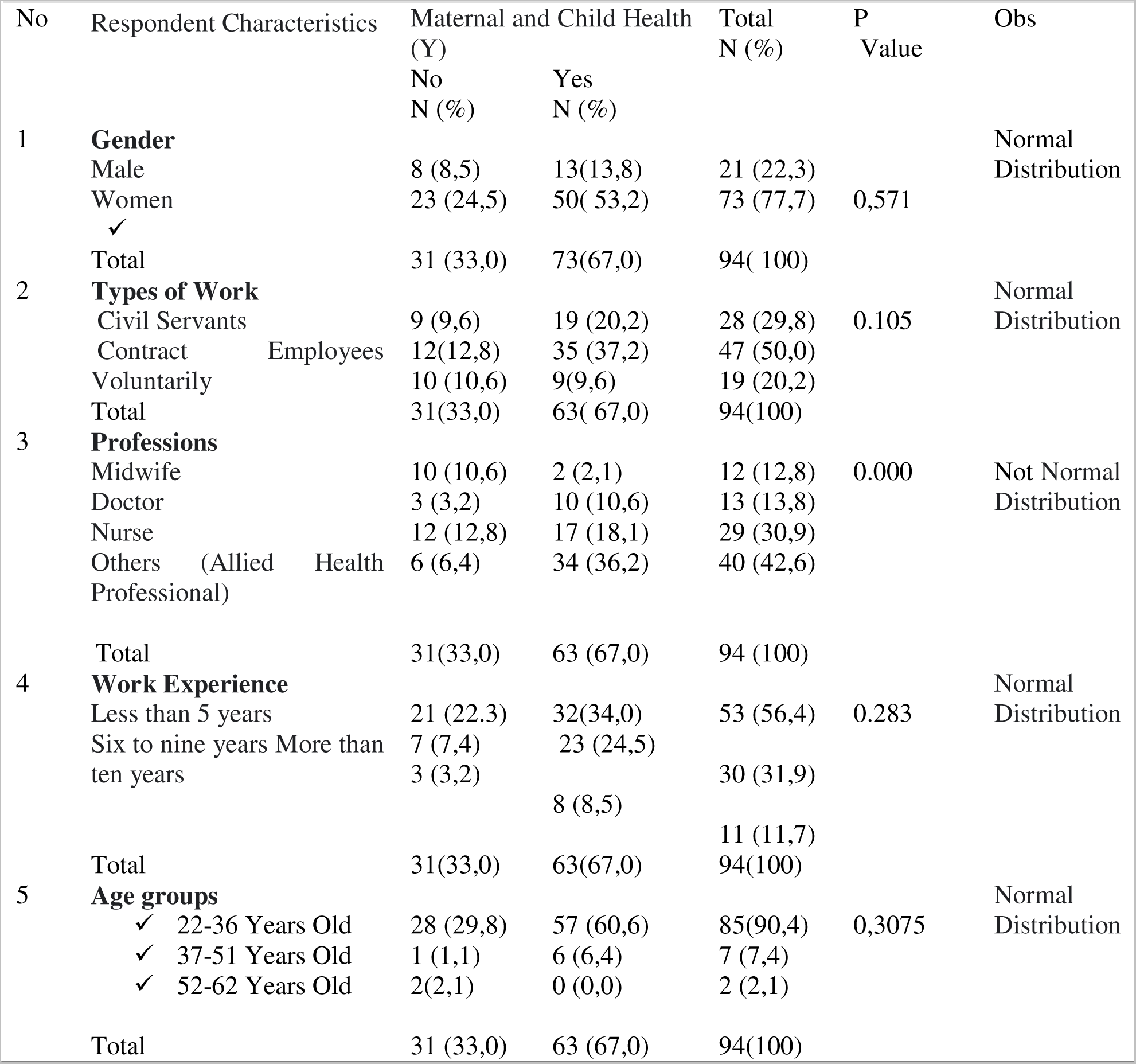
Data of Respondent Characteristics.

While, looking at the data on types of professions, it shows that the respondents composed of 12.8% midwives, 13.8% doctors, 30.9% nurses, and 42.6% Allied health professionals were tasked to deliver MCH services during the COVID-19 pandemic. However, about 33% of those health professionals working did not carry out MCH services during the pandemic. The most interruption was observed in Allied health for professionals (12.8%) and followed by midwives at 10.6%.

### The Association Between Health System Components and Disruption MCH Services during COVID-19 Pandemic

The Bivariate Analysis *(Crosstabulation)* of the association between health systems disruption and COVID-19 Pandemic was presented well in table-3. Each health system component was cross-tabulate using Bivariate Analysis to assess the association of its disruption caused by COVID-19 Pandemic.

**Table 3:**
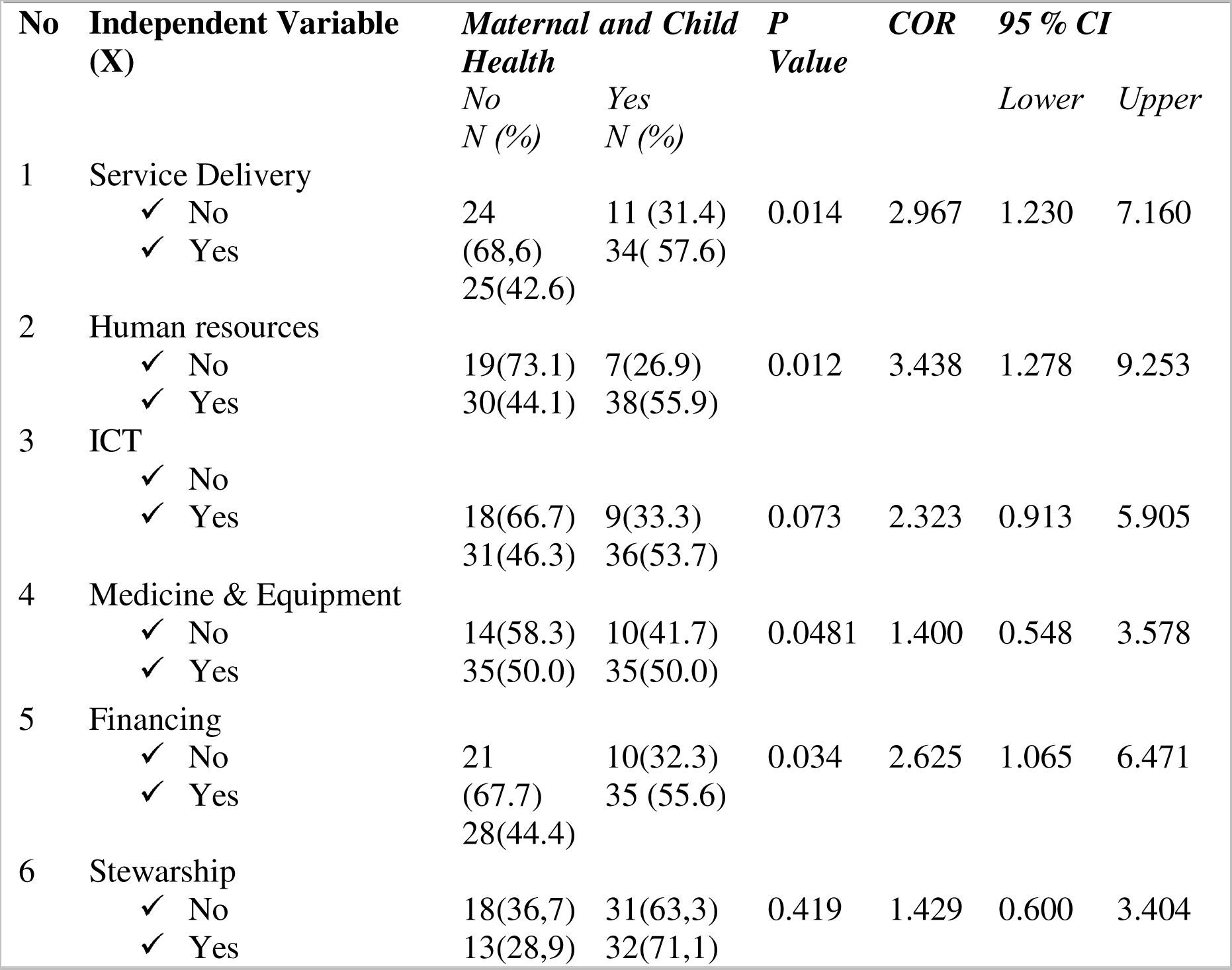
The Bivariate Analysis *(Crosstabulation*)

4.2.1 The Association between Services Delivery component and Disruption of MCH Services during COVID-19 Pandemic Table 3 shows that 68.6% of respondents stated that health services delivery of MCH care was disrupted during COVID-19 Pandemic.
4.2.2 The Association between Human Resource component and Disruption of MCH Services during COVID-19 Pandemic Table 3 shows that (73,1%) of respondents stated that on unavailability of some health professionals to provide MCH care during COVID-19 Pandemic.
4.2.3 The Association between ICT component and Disruption of MCH Services during COVID-19 Pandemic Table 3 shows that 66,7% of respondents stated that ICT component for MCH care was not utilized to disseminate information and consultation on MCH Care during COVID-19 Pandemic.
4.2.4 The Association between the availability of essential medicines and equipment component and Disruption of MCH Services during COVID-19 Pandemic Table 3 shows that 58.3 % of respondents stated that the unavailability of some essential medicines, equipment, PPE for MCH care during COVID-19 Pandemic.
4.2.5 The Association between the availability of financing component and Disruption of MCH Services during COVID-19 Pandemic Table 3 shows that 67.7 % of respondents stated that the less availability of finance for support MCH Care during COVID-19 Pandemic.
4.2.6 The Association between the stewardship component and Disruption of MCH Services during COVID-19 Pandemic Table 3 shows only 36.7 % of respondents stated that the stewardship provided to MHC services was interrupted was interrupted during COVID-19 Pandemic.

## DISCUSSION

This study has successfully collected frontline health care workers (HCWs) opinion regarding the System performance of MCH care in a primary health care facility during COVID-19 pandemic. This is the first study utilizing modified WHO Six building blocks for assessing health system performance [7] to evaluate the disruption of health system components of MCH Care in a community health center (CHC) during COVID-19 Pandemic in Timor-Leste. So far, most of the studies that evaluates MCH Care performance during COVID-19 Pandemic were focusing mostly on the service delivery component of the health care system. [12] [11] [3] [10] [13] [14]

The overall findings from this study reveals that all six health system components (blocks) for MCH care were disrupted with different degree of percentage during COVID-19 pandemic. This is very important information to help develop better system preparedness and better response to pregnant women and newborns in the future pandemic. [9] [8] [10] Relying on information from service delivery (one component) only, may mislead the policy makers to prepare appropriate plans for MCH care in at Primary Health Care Institution (PHCI). A health system-based approach at PHCI must be comprehensive both in terms of treating COVID-19 and non-COVID-19 patients. [25] G

### Provision Of MCH Services

High percentage (68.6%) of respondents said they did not carry out regular health services for mothers and children during the COVID-19 Pandemic. This finding in line with the cohort data on the reduction of ANC1, ANC4, Facility Delivery, and postnatal care (PNC) in 2020 during COVID-19 Pandemic compared to 2019 in Comoro-CHC. [26] The situation was worsening during 2021 due to community transmission of Delta and Omicron variant in the country caused many deaths and hospitalization. [21]This finding of maternal and child health services disruption was similar with research findings from many developing countries in Asia and Africa region. [3] [10] [11] [12] [13]. A rapid review carried out on PNC services availability and utilization during the COVID-19 era in sub-Saharan Africa revealed that there were significant declines in the availability and utilization services during and after the COVID-19 lockdown. [14].

The interruption of services delivery in Timor-Leste may be caused by many reasons including lockdown restriction and mandatory stay home order implemented in the country from April 2020 to November 2021. [20] [21] Interviews with healthcare providers and clients from study in Ethiopia highlighted several barriers to service utilization during COVID-19, including fear of disease transmission, economic hardship, and transport service disruptions and restrictions. [11] Kapoor and collaegues.2022, in their study found a causal link between restrictions and health service utilization and call for policy makers in low- and middle-income countries to carefully consider the trade-offs of strict lockdowns during future COVID-19 waves or future pandemics. [27] Study from Tanzania suggest that a part erratic political response, it was rather fear of the pandemic itself and diversion of resources to control COVID-19, that may have contributed most to lower the utilization of mother and child services. [28]

### The Availability of Health Care Workers

During the pandemic, health care workers (HCWs) have been in the frontline of health system response. This requires them to acquire knowledge and protection through personnel protective equipment (PPE) and Immunization in order to delivery health services. We found high percentage (73.1%) of frontline health workers interviewed in this study said they did not carry out work as a health profession regularly. One of the reason among others is because they were infected with COVID-19, especially during the second and third wave caused by Delta and Omicron variant. [21] During the periods of COVID-19 pandemic many health care professionals were trained and relocated to COVID-19 treatment centers to provide care to COVID-19 patients and increase their chance to get infected. [20] [21] In Timor-Leste, HCWs were at the high risks of getting infected compare to the general population. Evidence from seroprevalence study among HCWs indicating a significant sub-clinical infection among HCWs early in the local outbreak. [29].

Global studies have documented on the morbidity and mortality the frontline health care workers during COVID-19 Pandemic. Various reasons were identified contributing to the cause of Morbidity and Mortality of HCS were including lack of PPE, Not received Immunization, lack of supporting facilities and infrastructure, excessive working hours, and mental health problems [30] [31] [32] [33], [34], [35] [36], [37], [38] [39] There were also other reasons that caused interruption of HCWs from health facilities during COVID-19 Pandemics. They are including factors such as less motivated than usual to work, ration the use of PPE, scared of being infected, concerned for family members get infected, and anxiety. [33] [35] Therefore, It is important for the future pandemic to equip these frontline health workers with knowledge, equipment, proper infrastructure, incentive and moral support to keep them working and serving patients both COVID-19 and non Covid-19.

### The availability of MCH Medical products and equipment

During the pandemic, the most attention and resources were diverting towards preventing, diagnostic and treating COVID-19. Despite, MOH call for ensuring the delivery of essentials services through-out the country, but many other these services were disrupted. [20] About 58.3% HCWs interviewed in this study answer that there were stock-out of essential medicines and health products for the MCH service program at this Community Health Center during the COVID-19 pandemic. The COVID-19 pandemic reduces the capacity of SAMES to procure health products due to country lockdown and supplier issues. Despite alternative mechanism was sought to UN procurement system to purchase COVID-19 health products and essential medicine, [20] it’s can only provide short-term solution for chronic and ongoing problems. The stock-out of essential medicines and health products has always been issues for Timor-Leste for decades. [40] [20] Various factors including change management, prescriptions problems, and no pharmaceuticals industries in the country were contributing to ongoing stock-out of essential medicines and health products in Timor-Leste.

The issues of stocks-out of health products and equipment were also documented from other developing countries. In India and Nepal also the pandemic has decreased availability of certain medications necessary for pregnant women, and decreased access to contraceptives for others. [41] In Ethiopia, study from hospital settings reveal that the availability of key essential medicines for maternal and child health in the study area was low. The overall stock-out situation of MCH products has worsened during COVID-19 compared to pre-COVID-19 pandemic. [42]

Therefore, to ensure that stock outs of medicines and health products do not occur in the future, it is necessary to provide capacity development regarding supply chain management training to staff holding pharmaceutical units. Delivery of drugs and other products between consumers should be through rationalized prescriptions by medical personnel, so that recommended interventions can be used judiciously. [25]

### Information, Communication, and Technology Used for MCH Services

Despite, there was huge investment and utilization of ICT for COVID-19 during the pandemic in Timor-Leste. [20] [43] About 66.7% of HCWs interviewed in study said there was not available information, communication and utilization of technology for MCH care during COVID-19 Pandemic. This is true because during the pandemic most of the information, communication, and technology was directing towards on prevention of COVID-19 and to provide the right information on adverse effects of AstraZeneca that were emerged during the early deployment of the Vaccine in Timor-Leste. [43] Before COVID-19 pandemic, communication and information a [20] activities on MCH were regularly implemented are in all health facilities and through-out communities via SISCa, SNF and Villages Midwives. The use of digital health applications has increasingly become an important and strengthened during COVID-19 Pandemic. Despite huge challenges of mobile cellular networks, slow internet, and lack of digital infrastructure. Before the pandemic, there was one digital health intervention in operation – *Liga Inan*, which connects pregnant mothers and their health providers through mobile phones. [44] The *Liga-Inan* program was one of the reliable digital tools that continue provide pregnancy related information to all pregnant mother and child during the COVID-19 pandemic in Timor-Leste.

The Utilization of Digital Technology for sharing information and medical training such telehealth was increasingly utilizing in many global settings. Enhancing primary healthcare, use of telehealth, extended prescription, and public communication were countermeasures undertaken by China during the rapid rise period. [45] Uptake of telehealth and a shift from in-person care has been a major contributor to maintaining pregnancy care during pandemic restrictions in, New South Wales, Australia. [46]Researcher from Africa call for the use of telemedicine needs to be improved in African countries to allow for the continued provision of health services during pandemics. [47] Globally, Various (IEC) materials have been produced to raise awareness regarding protection of mother and child during COVID-19 through professional associations and made available free only [8] However, due to internet connecting challenges, these free materials were often lacking the lower level health facilities especially in the post-conflict and developing countries. The utilization of various mechanism of Internet of Things (IOT) and digitalization of health that was previously utilized for COVID-19 in Timor-Leste, such as WhatsApp Group (WG), Zoom, Google meet, COVID-19 Dashboard, and online call, [20], shall be further expanding to cover other essential services including MCH, continue post-pandemic and strengthening for future pandemic.

### Leadership and Financing MCH Services

Effective leadership and governance ensure the existence of strategic policy frameworks, effective oversight and coalition building, provision of appropriate incentives, and attention to system design, and accountability. [7]The majority (71.1%) of HCWs interviewed for this study respond on the good leadership provided for MCH during COVID-19 Pandemic. As argued by Joshua F and collegues, 2023, the eeffective national-level coordination of the response to the pandemic was important to ensure a cross-sectoral government approach, and because of the imperative of coordinating inputs from multiple development partners operating within Timor-Leste. [43] Early in the pandemic, the new governance structure of the Integrated Centre for Crisis Management (Centro Integrado de Gestao de Crises (CIGC)) was established under the Prime Minister’s office to provide leadership and governance in dealing with the COVID-19. [20]). Leadership in a health system includes setting priorities and the overall vision and direction A good leader will also ensure that policies and regulations are in place for effective and safe service delivery, and that appropriate mechanisms are in place to ensure accountability. Stewardship of the health care sector includes coordination between service delivery agencies, donors and other ministries. [48] Ministry of Health, 2020 issues special instruction for pregnant women or mother infected with COVID-19 to be treated with intensive care and also instruction to treat pregnant women and mother without infection. [49]The Pillar-9 under CIGC -Task Force was responsible to ensure the continuity delivery of essential health care services, including MCH services through-out the country. [20]

A good health financing system raises adequate funds for health, protects people from financial catastrophe, allocates resources, and purchases good and services in ways that improve quality, equity, and efficiency. [7] Funding should be made available for a quick and effective response to emergencies, requiring a supportive flexible public financial management system. Especially for the outpatient care, can also minimize the potential infection in health facilities. [50] This was not the case in Timor Leste during experiencing COVID-19 Pandemic. About 67.7 % of HCWs interviewed in this study stated the unavailability of finance resources to support MCH Care during COVID-19 Pandemic. In contrast, in respond to the pandemic, Parliament of Timor Leste allow the funding from the Petroleum Fund (a national wealth fund used for emergency purposes) in used for an extraordinary transfer to the state budget to fund COVID-19 health services and to maintaining essential health services. [21] Similarly, In Asia and the Pacific, COVID-19 services in most countries have been funded by government budgets and public insurance systems. In China, Indonesia, Malaysia, Mongolia, and Vietnam, for example, government budgets were used for both regular and emergency purposes. [50] Study from Nigeria report on the huge out-of-pocket expenses, and the least responsive of the COVID-19-related commodity procurement to the needs of those most in need of care and support.. [51]

### Study Limitation and Strength

Limitation: This results study present here is based only on the interviewed of HCWs from Comoro CHC. Therefore, it does not represent situation of all health facilities in Dili municipalities and the whole country. By the nature of study (cross-sectional study/survey) is prone to selection biases, interview biases and interpretation biases.

Strength: This is the first study that utilizing WHO six building Blocks to assess the system performance of MCH in a primary health care facility in Timor Leste. The study interviewed mostly COVID-19 frontline health care workers that directly working under the system during COVID-19 Pandemic.

Future research utilizing mix-methods combining quantitative interview with document review and field observation against the six-health system block in all community health centers in Dili or other municipalities, will provide better picture on the health system disruption for MCH care in Timor-Leste.

## CONCLUSION

This study has successfully collected frontline health care workers (HCWs) opinion regarding the System performance of MCH care in a primary health care facility during COVID-19 pandemic. This is the first study utilizing modified WHO Six building blocks for assess the disruption of health system components of MCH Care in a community health center (CHC) during COVID-19 Pandemic in Timor Leste. The overall findings from this study reveals that all six health system components (blocks) for MCH care were disrupted with different degree of percentage during COVID-19 pandemic. This is very important information to help develop better system preparedness and better response to pregnant women and newborns in the future pandemic. Relying on information from service delivery (one component) only, may mislead the policy makers to prepare appropriate plans for MCH care in at Primary Health Care Institution (PHCI) in the events of Pandemic. For future emergency plan, the attention must be given to all six-health system blocks to guarantee continue delivery of MCH Care in a primary health care clinic, in Timor-Leste and other similar settings.

## ACKNOLEDGEMENT

We thank to the Director Executive of Instituto Nacional de Saúde (INS) MoH and Rector of Universidade da Paz, Timor-Leste, were provided motivation support during the study.

## AUTHOR CONTRIBUTION

Each author was contributed in this study for conception, design, data collection, data analysis and interpretation, result writing and manuscript drafting.

## CONFLIC OF INTERES

No any conflict of interest in this study.

## ETHICAL CONCIDERATION

The Instituto Nacional de Saúde (INS) MoH through Ethical committee was provided approval letter for this study conducting.

## Data Availability

All data produced in the present study are available upon reasonable request to the authors

